# Pubertal timing in boys and girls born to mothers with gestational diabetes mellitus: a systematic review

**DOI:** 10.1101/2020.03.20.20039685

**Authors:** Anuradhaa Subramanian, Jan Idkowiak, Konstantinos A. Toulis, Shakila Thangaratinam, Wiebke Arlt, Krishnarajah Nirantharakumar

**Affiliations:** Institute of Applied Health Research, University of Birmingham, B15 2TT, United Kingdom; Centre for Endocrinology, Diabetes and Metabolism, Birmingham Health Partners, University Hospitals Birmingham NHS Foundation Trust, Birmingham, B15 2GW, United Kingdom; Institute of Metabolism and Systems Research, University of Birmingham, B15 2TT, United Kingdom; Department of Paediatric Endocrinology and Diabetes, Birmingham Children’s Hospital, Birmingham Women’s and Children’s Hospital NHS Foundation Trust, Birmingham, B4 6NH United Kingdom

**Keywords:** Adrenarche, pubarche, gonadarche, menarche, offspring health, metabolic risk

## Abstract

**Context:** The incidence of gestational diabetes mellitus (GDM) has been on the rise, driven by maternal obesity. In parallel, pubertal tempo has increased in the general population, driven by childhood obesity.

**Ojective:** To evaluate the available evidence on pubertal timing of boys and girls born to mothers with GDM.

**Data Sources:** We searched MEDLINE, EMBASE, CINAHL Plus, Cochrane library and grey literature for observational studies up to October 2019.

**Study selection and extraction:** Two reviewers independently selected studies, collected data and appraised study quality. Results were tabulated and narratively described as reported in the primary studies.

**Results:** Seven articles (six for girls and four for boys) were included. Study quality score was mostly moderate (ranging from 4 to 10 out of 11). In girls born to mothers with GDM, estimates suggest earlier timing of pubarche, thelarche and menarche although for each of these outcomes only one study each showed a statistically significant association. In boys, there was some association between maternal GDM and earlier pubarche, but inconsistency in the direction of shift of age at onset of genital and testicular development and first ejaculation. Only a single study analysed growth patterns in children of mothers with GDM, describing a 3-month advancement in the age of attainment of peak height velocity and a slight increase in pubertal tempo.

**Conclusions:** Pubertal timing may be influenced by the presence of maternal GDM, though current evidence is sparse and of limited quality. Prospective cohort studies should be conducted, ideally coupled with objective biochemical tests.

## INTRODUCTION

Puberty marks an important period in the dynamics of childhood development characterised by fundamental physical, cognitive and psychological transformation. The attainment of adult-like secondary sexual characteristics, rapid growth, changes of body composition and achieving fertility are the main physical outcomes of puberty. As a consequence of the maturation of the hypothalamic-pituitary-gonadal axis with subsequent incremental, finely-orchestrated gonadal sex steroid production, typical physical changes occur in a successive fashion. In girls, this usually starts with thelarche (onset of breast development) and pubarche (appearance of pubic hair), followed by a peak growth spurt culminating in menarche (first menstruation) (1). In boys, testicular enlargement and pubarche are the first physical signs of puberty followed by peak growth spurt and spermarche (development of sperm) with the occurrence of the first ejaculation.

A secular trend of advancement in pubertal timing along with a steep decline in the age of menarche from 17 to 13 years has been recognized between 19^th^ and 21^st^ century (1-3). Consequently, increasing numbers of children are diagnosed with central precocious puberty (1,2), defined as the onset of gonadarche before the age of 8 years in girls and 9 years in boys, based upon pubertal staging performed in the 1960s (4,5). Compared to peers who mature on-time or later, early developers are more likely to experience psychological distress and social isolation, potentially leading to detrimental outcomes such as poor academic performance, depression, substance abuse, eating disorder, disturbed body image and risky sexual behavior (6,7). Early pubertal timing also has an adverse impact on adult metabolic health including increased risk of diabetes and other cardio-vascular morbidity (6-8).

Risk factors for early puberty are considered to be multifactorial and may be seen as the effect of factors influencing the maturation of the hypothalamic GnRH pulse generator. These include predisposing genetic factors, intrauterine environment, and endocrine disrupting chemicals, and, first and foremost, abundance of nutrients and childhood obesity (9). Similar to the trend towards earlier pubertal timing driven by childhood obesity, the incidence of gestational diabetes mellitus (GDM) driven by maternal obesity has also been on the rise; in the last decade, the incidence of GDM has doubled from 2.7% to 5.6% and is predicted to further increase (10).

The effect of maternal GDM on pre-pubertal health outcomes in the offspring has been evaluated by a limited number of observational studies, but evidence on the effect of GDM on sexual maturation and pubertal timing is scarce and conflicting. Due to the complexity in the conceptualization of pubertal timing and its clinical assessment and the significant heterogeneity among the studies exploring the relationship between maternal GDM and central precocious puberty, a causal relationship has not been clearly established yet. If confirmed, such a link could drive a transgenerational continuum and, thereby, metabolic morbidity associated with both conditions. Here, we have undertaken a systematic appraisal of the available evidence on pubertal timing in children born to mothers with GDM.

## METHODS

### Searches

We carried out a systematic literature review search initially in March 2019, with the search rerun in October 2019 to retrieve any additional studies before final synthesis of results. Databases included: (1) Electronic bibliographic databases (MEDLINE, EMBASE, CINAHL

Plus, Cochrane library), (2) Google Scholar™ search and experts contact to obtain relevant grey literature, and (3) citations tracked from the screened articles to identify further relevant studies. The search strategy was constructed with the help of a medical librarian combining natural and structured language terms (MESH and Emtree). Terms relating to “gestational diabetes” was combined with an ‘AND’ Boolean operator to “puberty”, “pubarche”, “thelarche”, “menarche”, “tanner staging”, “spermearche” and “growth”. A list of search terms is provided in Supplementary Table 1.

**Table 1.** Characteristics of the included studies.

| No. | Study | Location | Study database | GDM diagnostic criteria | Age of mothers | Offspring sex | Sample Size |  | Outcomes considered |
| --- | --- | --- | --- | --- | --- | --- | --- | --- | --- |
|  |  |  |  |  |  |  | GDM | Controls |  |
| 1 | D'Aloisio et al. 2013 (24) | USA and Puerto Rico | Sister Study | Self reports from family history questionnaires completed by the offspring | N (%) †<br>< 20 years : 1,537 (5%)<br>20-34 years: 2,5191 (76%)<br>>=35 years: 6,380 (19%) | Girls | 178 | 30,169 | Adjusted RR for age at menarche: ≤ 10, 11, 12-13 (reference age group), 14 and ≥ 15 years |
| 2 | Grunnet et al. 2017 (25) | Denmark | Danish National Birth Cohort (DNBC) | Danish National Patient Registry criteria* ± self reports | NR | Girls | 238 | 256 | Age-adjusted OR for ≥ Tanner B2 |
|  |  |  |  |  |  |  | 221 | 237 | Age-adjusted OR for ≥ Tanner PH2 |
|  |  |  |  |  | NR | Boys | 192 | 182 | Age-adjusted OR for testicular volume ≥ 4 mL |
|  |  |  |  |  |  |  | 206 | 203 | Age-adjusted OR for ≥ Tanner PH2 |
|  |  |  |  |  |  |  | 193 | 176 | Age-adjusted OR for ≥ Tanner G2 |
| 3 | Hockett et al. 2019 (26) | USA | Kaiser Permanente of Colorado Health Plan (KPCO) linked to Exploring Perinatal Outcomes among Children (EPOCH) study | as recommended by ational Diabetes Data Group (NDDG) ** | NR | Girls | 34<sup>§</sup> | 174 | Age at PHV stratified by exposure status.<br>PHV stratified by exposure status.<br>And beta coefficients and p-value from accelerated failure time model and p-value. |
|  |  |  |  |  | NR | Boys | 43<sup>§</sup> | 166 | Age at PHV stratified by exposure status.<br>PHV stratified by exposure status.<br>and beta coefficients and p-value from accelerated failure time model and p-value |
| 4 | Kubo et al. 2015 (27) | USA | Kaiser Permanente Northern California (KPNC) linked to Cohort Study of Young Girls' | Carpenter and Coustan criteria*** | Mean (SD)<br>GDM – 33.0 (5.0)<br>Control – 32.6 (5.9) | Girls | 27 | 390 | Adjusted HR:<br>≥ Tanner B2<br>≥ Tanner PH2 |
|  |  |  | Nutrition,<br>Environment<br>and Transitions<br>(CYGNET) |  |  |  |  |  |  |
| 5 | Kubo et al.<br>2018 (28) | USA | Kaiser<br>Permanente<br>Northern<br>California<br>(KPNC) | Carpenter and<br>Coustan<br>criteria*** | Mean (SD) †<br>UW-27.9(5.2)<br>NW- 29.9(5.1)<br>OW- 30.5(5.2)<br>OB- 30.3(5.3) | Girls | 977 | 11,364 | Adjusted HR for ≥ Tanner B2 |
|  |  |  |  |  |  |  | 931 | 10,839 | Adjusted HR for ≥ Tanner PH2 |
| 6 | Lauridsen<br>et al.<br>2018 (29) | Denmark | The Danish<br>National Birth<br>Cohort (DNBC)<br>linked to the<br>Puberty Cohort | Danish National<br>Patient Registry*<br>+ self reports | Median (IQR)<br>GDM – 31.9 (31.4-32.5)<br>Control – 30.6 (30.5-30.7) | Girls | 122 | 15,320 | Adjusted mean monthly difference<br>for:<br>Tanner B2, B3, B4, B5<br>Tanner PH2, PH3, PH4, PH5<br>Menarche<br>Axillary hair<br>Acne |
|  |  |  |  |  |  | Boys | 130 |  | Adjusted mean monthly difference<br>for:<br>Tanner G2, G3, G4, G5<br>Tanner PH2, PH3, PH4, PH5<br>Spermarche<br>Voice break<br>Adult Voice<br>Axillary hair<br>Acne |
| 7 | Monteilh et<br>al. 2010<br>(30) | Avon,<br>England | Avon<br>Longitudinal<br>Study of Parents<br>and Children | Self reports | Mean (SD) †<br>29.0 (3.8) | Boys | 450 | 3,263 | Median age at transition to:<br>Tanner PH>1, PH>2 and PH>3<br>based on multi-stage modelling |

†, summarizes the cohort from the primary study [Potential contamination from mothers with pre-existing diabetes or patients with missing information on GDM];
§, 10% contamination of the GDM exposed cohort with exposure to type 1 diabetes;
\*, Danish National Patient Registry Criteria: GDM diagnosis is made based on WHO criteria (see below) and in some cases if two or more glucose values measured exceeded the mean ± 3 SD on a curve based on a group of 40 healthy, nonobese, nonpregnant Danish women without a family history of diabetes , WHO Criteria: fasting blood glucose > 7.0 mmol/L or random blood glucose > 11.1 mmol/L or 75-g oral glucose tolerance test at 2-hours > 11.1 mmol/L;
\*\*, NDDG criteria: 50-g oral glucose tolerance test at 1-hour ≥7.8 mmol/L followed by a 100-g oral glucose tolerance test at 3-hours > 7.8 mmol/L;
\*\*\*, Carpenter and Coustan Criteria: fasting blood glucose > 5.3 mmol/L or 100-g oral glucose tolerance test at 1-hour > 10 mmol/L, at 2-hours > 8.3 mmol/L, at 3-hours > 7.8 mmol/L
*Abbreviations:* NR, not reported; RR, Risk Ratio; OR, Odds Ratio; HR, Hazard Ratio; UW, underweight; NW, normal weight; OW, overweight; OB, obese;
PHV, peak height velocity

Records identified by the searches were independently screened by two reviewers (A.S. and J.I.) in the order of title, abstract and full text of the article. Articles were selected when they met the inclusion and exclusion criteria mentioned in the predefined protocol registered on PROSPERO [CRD42019150365]. In case of study selection disagreements, a third reviewer (K.N.) was consulted to reach consensus.

### Inclusion and Exclusion Criteria

We included observational studies – cohort, case-control and cross-sectional studies. Studies that considered multiple exposures or multiple outcomes were also included, if they studied the association between maternal GDM and pubertal timing in the offspring. Pubertal timing was allowed to be described by the timing of the following pubertal milestones according to Tanner (4,5): in girls, 1) pubic hair development/ pubarche (Tanner Stage: ≥PH2), 2) breast development/ thelarche (Tanner Stage: ≥B2), 3) menarche and 4) speed of pubertal growth as peak height velocity (PHV) and age at PHV; in boys, 1) pubic hair development/pubarche (Tanner Stage: ≥PH2), 2) testicular enlargement (testicular volume ≥4mL on either or both sides), 3) maturation of the external male genitalia (Tanner Stage: ≥G2), 4) spermarche and 5) PHV and age at PHV.

Studies were excluded if they were case studies, case series or commentary articles, qualitative studies without quantitative data on pubertal timing, studies reporting pubertal staging instead of pubertal timing disregarding chronological age, or studies conducted on non-human subjects.

### Data extraction and quality appraisal

The JBI data extraction form(11) was adapted based on the specifics of this review to create a template form in Microsoft Word® (Supplementary Table 2). The form mandated data on the following elements from the included studies: authors, study publication date, data source, study period, country and setting, sample size, GDM exposure ascertainment criteria, proportion of GDM exposed women who used insulin, offspring sex, outcome/s considered and details on analytical methods employed including the list of confounding variables considered.

**Table 2:** Evidence summary of the relationship between maternal GDM and pubertal development in their daughters identified by pubic hair development (n=4 studies), breast development (n=4), peak height velocity (n=1), menarche (n=2) and other pubertal changes (n=1)

| Outcome | Study ID | Outcome Metrics | Estimates |
| --- | --- | --- | --- |
| Pubic hair development/<br>Pubarche | Grunnet et al., 2017 (25) | N (%) of ≥ Tanner PH2 in GDM and controls<br>Age-adjusted OR (95% CI) of ≥ Tanner PH2 | GDM: 99 (44.8%)<br>Controls: 133 (56.1%)<br>OR:<br>adjusted: 1.51 (0.90,2.55); p=0.12 |
| | Kubo et al., 2018 (28) | Adjusted (for race/ethnicity, maternal age, education, parity, smoking during pregnancy and BMI) HR (95% CI) of ≥ Tanner PH2 | HR<br>adjusted: 1.04 (0.92,1.17) |
|  | Lauridsen et al., 2018 (29) | Mean age at Tanner PH2, PH3, PH4, PH5 in daughters of mothers without diabetes | Mean Age (years):<br>PH2: 11.3; PH3: 12.5; PH4: 13.4; PH5: 15.3 |
|  |  | Crude and adjusted (for maternal age at menarche, maternal age at birth, socioeconomic status, cohabitation, parity and maternal BMI) mean monthly difference in age (95% CI) at Tanner PH2, PH3, PH4, PH5 | Mean difference (months):<br>PH2:<br>crude -5.4; adjusted -4.8 (-7.7, -2.0)<br>PH3:<br>crude -3.1; adjusted -2.2 (-4.4, 0)<br>PH4:<br>crude -2.6; adjusted -1.6 (-4.8, 1.6)<br>PH5:<br>crude -7.8; adjusted -6.0 (-10.8, -1.2) |
| | Kubo et al., 2016 (27) | Unadjusted and adjusted (for race/ethnicity, household income and maternal age) hazard ratio (95% CI) of ≥ Tanner PH2<br>Similar estimates for the interaction between maternal pregravid BMI and GDM: | HR:<br>crude: 1.09 (0.71,1.70);<br>adjusted: 1.24 (0.79,1.94)<br>HR:<br>adjusted:<br>BMI<25 & no GDM: Reference category<br>BMI<25& GDM: 1.06 (0.48,2.36)<br>BMI≥25 & no GDM: 1.19 (0.90,1.56) |
| | | | BMI≥25 & GDM: 2.97 (1.52,5.83) |
| Breast Development/<br>Thelarche | Grunnet et al., 2017 (25) | N (%)≥ Tanner B2 in GDM cases and controls<br>Age adjusted OR (95% CI) of ≥ Tanner B2 | GDM: 141 (59.2%)<br>controls: 169 (66.0%)<br>OR:<br>adjusted: 1.99 (1.18,3.34) p=0.01<br>Estimate with additional adjustment for BMI shown in primary study's figure (Direction of effect remains but statistically NS) |
| | Kubo et al., 2018 (28) | Adjusted (for race/ethnicity, maternal age, education, parity, smoking during pregnancy and BMI) hazard ratio (95% CI) of ≥ Tanner B2 | HR:<br>adjusted: 1.06 (0.95,1.18) |
|  | Lauridsen et al., 2018 (29) | Mean age at Tanner stage B2, B3, B4, B5 in daughters of mothers without diabetes<br>Crude and adjusted (for maternal age at menarche, maternal age at birth, socioeconomic status, cohabitation, parity and maternal BMI) mean monthly difference in age (95% CI) at Tanner stage B2, B3, B4, B5 | Mean Age (years):<br>B2: 9.9 ; B3:11.6; B4:13.0; B5: 15.7<br>Mean difference (months):<br>B2:<br>crude -7.3;adjusted -4.6 (-10.1, 1.0)<br>B3:<br>crude -3.8;adjusted -1.9 (-5.0, 1.2)<br>B4:<br>crude -2.1;adjusted -0.5 (-3.2, 2.4)<br>B5:<br>crude -4.3;adjusted -1.8 (-7.9, 4.3) |
| | Kubo et al., 2016 (27) | Unadjusted and adjusted (for race/ethnicity, household income and maternal age) hazard ratio (95% CI) of breast development stage≥B2<br>Similar estimates for pregravid BMI # GDM interaction: | HR:<br>crude: 1.01 (0.65,1.57)<br>adjusted: 0.85 (0.54,1.35)<br>HR:<br>adjusted:<br>BMI<25#No GDM: Reference category<br>BMI<25#GDM: 1.22 (0.54,2.74)<br>BMI≥25#No GDM: 1.18 (0.88,1.57)<br>BMI≥25#GDM: 0.93 (0.47,1.85) |
| PHV and age | Hockett et al., 2019 | Age at PHV stratified by maternal GDM exposure status | Age at PHV (years) |
| at PHV | (26) | and beta coefficient for exposure to GDM in utero after adjusting for child's race/ethnicity<br>PHV among exposed and unexposed girls and boys and beta coefficient for exposure to GDM in utero after adjusting for child's race/ethnicity | GDM: 10.85<br>No GDM: 11.12<br>Beta co-efficient and p-value not reported [but figure shows overlapping confidence intervals of age at PHV between exposed and the unexposed]<br>PHV (cm/year):<br>GDM: 8.88<br>No GDM: 8.04<br>Beta Coefficient: 0.10; p<0.001 |
| Menarche | D'Aloisio et al., 2013 (24) | Relative Risk (95% CI) of earlier or later menarche in mothers with GDM compared to mothers without GDM after adjustment for birth decade, race/ethnicity, childhood family income, interaction between birth decade and race/ethnicity | RR:<br>adjusted:<br>≤ 10 years: 0.98 (0.53,1.84)<br>11 years: 0.99 (0.63,1.54)<br>12-13 years: Reference category<br>14 years : 0.77 (0.48,1.25)<br>≥ 15 years: 0.98 (0.60,1.60) |
|  | Lauridsen et al., 2018 (29) | Mean age at menarche in daughters of mothers without diabetes<br>Crude and adjusted (for maternal age at menarche, maternal age at birth, socioeconomic status, cohabitation, parity and maternal BMI) mean monthly difference in age (95% CI) at menarche | Mean Age(years): 13.0<br>Mean difference (months):<br>crude -4.1; adjusted -2.5 (-4.9, 0.0) |
| Other pubertal outcomes | Lauridsen et al., 2018 (29) | Crude and adjusted (for maternal age at menarche, maternal age at birth, socioeconomic status, cohabitation, parity and maternal BMI) mean monthly difference in age (95% CI) at development of axillary hair and acne | Mean difference (months):<br>AH:<br>crude -4.4; adjusted -3.6 (-7.3, 0.1)<br>Acne:<br>Crude -3.8; adjusted -2.6 (-6.8, 1.6) |

*Abbreviations:* RR, Risk Ratio; OR, Odds Ratio; HR, Hazard Ratio; NS, not significant; PH, Pubic Hair; B, Breast development; PHV, peak height velocity; NS, not significant; AH, Axillary Hair

An adapted version of the Newcastle-Ottawa critical appraisal checklist (12) was used to evaluate the quality of each of the included studies and individual studies were graded as low or high risk for each of the checklist questions (Template form is provided in Supplementary Table 3). Elements employed in appraising the internal validity of the included studies included: potential selection bias, objective GDM diagnosis and pubertal staging measurement, capture of and adjustment for confounding variables, appropriateness of statistical analysis employed, sufficient follow-up period and characteristics of patients lost to follow-up. Representativeness of the study population was also discussed to assess external validity.

**Table 3:** Evidence summary: The relationship between maternal GDM and pubertal development in their sons identified by pubic hair development (n=3 studies), testicular development (n=2), genital development (n=2), peak height velocity (n=1), spermearche (n=1) and other pubertal changes (n=1).

| Outcome | Study ID | Outcome Metrics | Estimates |
| --- | --- | --- | --- |
| Pubic hair development/<br>Pubarche | Grunnet et al., 2017 (25) | N(%) of ≥ Tanner stage PH2 in GDM cases and controls<br>Age adjusted OR (95% CI) of ≥ Tanner stage PH2 | GDM: 50 (24.3%)<br>Controls: 60 (29.6%)<br>OR:<br>adjusted: 1.74 (0.92,3.28) p=0.09<br>Estimate with additional adjustment for BMI shown in primary study's figure (Direction of effect remains and still statistically NS) |
|  | Lauridsen et al., 2018 (29) | Mean age at Tanner stage PH2, PH3, PH4,PH5 in sons of mothers without diabetes<br>Crude and adjusted (for maternal age at menarche, maternal age at birth, socioeconomic status, cohabitation, parity and maternal BMI) mean monthly difference in age (95% CI) at Tanner stage PH2, PH3, PH4, PH5 | Mean Age (years)<br>PH2: 11.3; PH3: 12.7; PH4: 13.5; PH5:14.7<br>Mean difference (months):<br>PH2:<br>crude -1.9; adjusted -1.4 (-5.3, 2.4)<br>PH3:<br>crude -1.9; adjusted -1.3 (-4.6, 1.9)<br>PH4:<br>crude -1.7; adjusted -0.8 (-3.4, 1.6)<br>PH:<br>crude -2.6; adjusted -1.7 (-4.7, 1.3) |
| | Monteilh et al., 2010 (30) | Multistage modelling:<br>Median age at transition (95% CI) to $>$Tanner stage PH1, $>$Tanner stage PH2, $>$Tanner stage PH3 | PH$>$1:<br>GDM was not a covariate in the combined multivariate model level due to statistical insignificance at the univariate analysis or restricted combined model level<br>PH$>$2 :<br>In the model with offspring BMI at age 8, |
|  |  |  | Median age at PH>2 (years): 12.8 (12.7-12.8)<br>Median age at PH>2 among sons of GDM mothers (years): 12.6 (12.4-12.7); p=0.03<br>In the model with offspring height and weight at age 8,<br>Median age at PH>2 (years) :13.0 (12.8-13.1)<br>Median age at PH>2 among sons of GDM mothers (years): 12.8 (12.6-13.0);<br>p=0.05<br>PH>3:<br>GDM was not a covariate in the combined multivariate model level due to statistical insignificance at the univariate analysis or restricted combined model level |
| Testicular development | Grunnet et al. 2017 (25) | N(%) of testicular volume $\geq 4$ mL in GDM cases and controls<br>Age adjusted OR (95% CI) of testicular volume $\geq 4$ mL | GDM: 143 (74.5%)<br>No GDM: 156 (85.7%)<br>OR:<br>Adjusted: 0.77 (0.42-1.41); p=0.40<br>Estimate with additional adjustment for BMI shown in primary study's figure (Direction of effect remains and still statistically NS) |
| Genital Development | Grunnet et al., 2017 (25) | N(%) of ≥ Tanner stage G2 in GDM cases and controls<br>Age adjusted OR (95% CI) of ≥ Tanner stage G2 | GDM: 63 (32.6%)<br>No GDM: 66 (37.5%)<br>OR:<br>Adjusted: 1.24 (0.72,2.14) p=0.45<br>Estimate with additional adjustment for BMI shown in primary study's figure (Direction of effect remains and still statistically NS) |
|  | Lauridsen et al., 2018 (29) | Mean age at Tanner stage G2, G3, G4, G5 in sons of mothers without diabetes | Mean Age (years):<br>G2:10.9; G3:12.5; G4: 13.6; G5:15.5<br>Mean difference (months): |
|  |  | Crude and adjusted (for maternal age at menarche, maternal age at birth, socioeconomic status, cohabitation, parity and maternal BMI) mean monthly difference in age (95% CI) at Tanner stage G2, G3, G4, G5 | G2:<br>crude -0.4; adjusted 0.0 (-3.8, 3.8)<br>G3:<br>crude 1.1; adjusted 1.4 (-1.9, 4.9)<br>G4:<br>crude -0.2; adjusted 0.5 (-2.5, 3.5)<br>G5:<br>crude 2.2; adjusted: 2.6 (-2.2, 7.4) |
| PHV and age at PHV | Hockett et al., 2019 (26) | Age at PHV stratified by exposure status and beta coefficient for exposure to GDM in utero after adjusting for child's race/ethnicity<br>PHV among exposed and unexposed girls and boys and beta coefficient for exposure to GDM in utero after adjusting for child's race/ethnicity | Age at PHV (years)<br>GDM: 12.68<br>No GDM: 12.92<br>Beta co-efficient and p-value not reported [but figure shows overlapping confidence intervals of age at PHV between exposed and the unexposed]<br>PHV (cm/year):<br>GDM: 9.65<br>No GDM: 9.28<br>Beta Coefficient : 0.04; p<0.001 |
| Spermarche | Lauridsen et al., 2018 (29) | Mean age at spermarche in sons of mothers without diabetes<br>Crude and adjusted (for maternal age at menarche, maternal age at birth, socioeconomic status, cohabitation, parity and maternal BMI) mean monthly difference in age (95% CI) at spermarche | Mean Age (years): 13.4<br>Mean difference (months):<br>crude 0; adjusted 0.7 (-2.9, 4.3) |
| Other pubertal outcomes | Lauridsen et al. 2018 (29) | Mean age at VB, AV, AH, acne in sons of mothers without diabetes<br>Crude and adjusted (for maternal age at menarche, maternal age at birth, socioeconomic status, cohabitation, parity and maternal BMI) mean monthly difference in age (95% CI) at voice break, adult voice, development of axillary hair and acne | Mean Age (years)<br>VB:13.0; AV: 15.0; AH:13.3; Acne:12.2<br>Mean difference (months):<br>VB:<br>crude -1.8;adjusted: -0.8 (-4.6, 2.8)<br>AV:<br>crude:-3.2; adjusted: -2.5 (-8.4, 3.4)<br>AH:<br>crude -4.3;adjusted: -2.9 (-7.4, 1.8)<br>Acne:<br>crude: 0.8; adjusted: 1.8 (-2.1, 5.7) |

*Abbreviations:* RR, Risk Ratio; OR, Odds Ratio; HR, Hazard Ratio; NS, not significant; Tanner stage PH, Tanner Stage Pubic Hair; Tanner Stage G, Tanner Stage Genital Development; PHV, peak height velocity; VB, Voice Break; AV, Adult Voice; AH, Axillary Hair

Data extraction and quality appraisal forms were pilot-tested with one of the included studies at the protocol-writing stage. Data extraction and quality appraisal were performed by two independent reviewers (A.S. and J.I.) and in case of disparities, a third reviewer (K.N.) was contacted to settle differences.

Findings of this review are reported in accordance with PRISMA guidelines (Supplementary Table 4)

## RESULTS

### Literature search results

We identified 305 studies through electronic database searches, including 57 duplicates (**Fig. 1**). Of the remaining 248, 230 were not relevant to the research question and were excluded on the basis of title and abstract, leaving 18 studies for full-text assessment. Eleven articles were excluded at this stage: four articles were conference proceedings, oral presentations or commentary articles (13-16); two articles did not include any of the outcomes we were interested in (17,18); one article did not analyse GDM as a predictor for pubertal timing due to an insufficient number of subjects with GDM (19); two articles did not provide a comparator cohort (20,21); two articles only reported Tanner stage at baseline and did not consider age/timing of puberty (22,23). The seven remaining studies were included in the review (**Fig. 1**).

**Figure 1.**
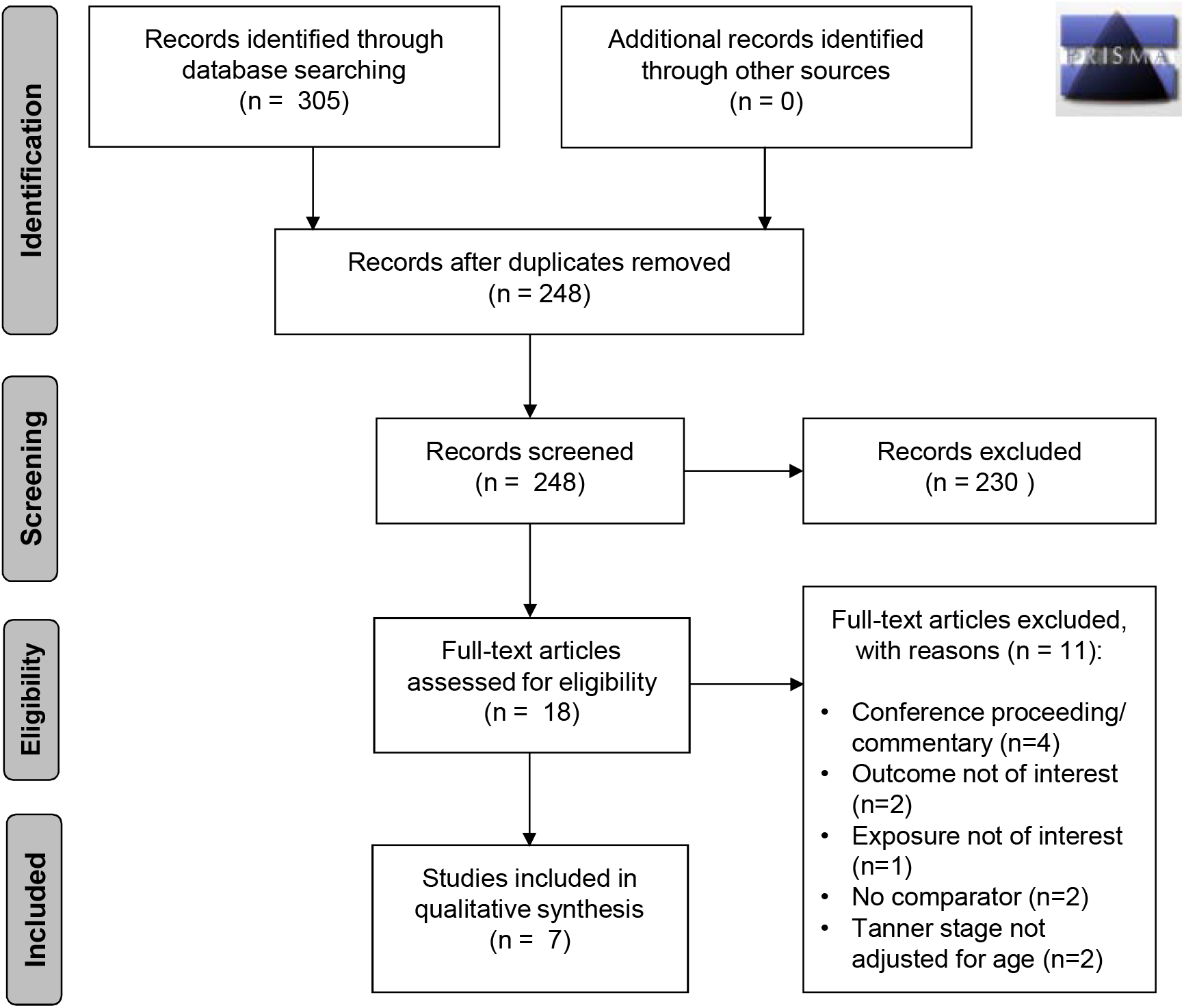
PRISMA Flowchart.

### Study Characteristics

The seven primary studies included in this review (24-30) are described in **Table 1**. Four studies were conducted in the USA (24,26-28), two in Denmark (25,29) and one in England (30). The populations studied were predominantly Caucasian. Four studies had comparable primary objectives to our review question,(26-29) two studies looked at multiple predictors of pubertal timing (24,30), and one looked at multiple developmental outcomes in the offspring of mothers with GDM including pubertal timing (25).

Three of the included studies focussed only on the pubertal timing in girls (24,27,28), one study focussed only on the pubertal timing in boys (30) and three studies reported outcomes for both boys and girls (25,26,29). All of the studies stratified their estimates by offspring sex.

Two pairs of the included articles derived their study sample from the same pregnancy cohorts and thus had the potential for overlapping populations [Danish National Birth Cohort (DNBC) [(25,29]) and Kaiser Permanante Northern California (KPNC) [(27,28)].

Sample size ranged widely both between and within studies when considering multiple outcomes (**Table 1**): D’Aloissio et al. included 33,501 daughters with information on age at menarche; 178 of them self-reported positive maternal GDM status through telephone contact with their mothers (24). Grunnet et al. considered multiple outcomes: breast and pubic hair development in 494 and 458 girls, respectively, and testicular volume, pubic hair, and genital developmental stage in 374, 409 and 369 boys, respectively (25). Hockett et al. included 208 girls and 209 boys with anthropometric records to calculate peak height velocity; 34 girls and 43 boys had positive maternal GDM status (26). Two studies that used the same cohort (KPNC) and considered the same outcomes varied with regard to the maternal sample size [417 and 12341] (27,28). Lauridsen et al. included 122 and 130 girls and boys with positive maternal GDM exposure status (29), while Monteilh et al. included 450 boys with positive maternal GDM exposure status (30).

### Quality Evaluation

The risk of bias based on the review question-adapted Newcastle Ottawa critical appraisal checklist is summarized for the seven included studies in **Fig. 2**. All populations studied were reasonably representative of their respective country’s general practice or hospital setting except for the study by D’Aloisio et al. (24), who had restricted inclusion to pregnant women at risk of breast cancer. Exposure information regarding GDM status was obtained from pregnancy registries in five studies (25-29), two of those studies also considered self-reports (25,29). However, for the remaining two studies (24,30), GDM status was only self-reported, indicating high risk of recall or misclassification bias. Studies based on KPNC cohorts mentioned using Carpenter and Coustan thresholds for GDM diagnosis. Variation was observed in the covariates considered, with race/ethnicity and socio-economic status representing the most popular confounders considered in the association between maternal GDM and pubertal timing in the offspring.

**Figure 2.**
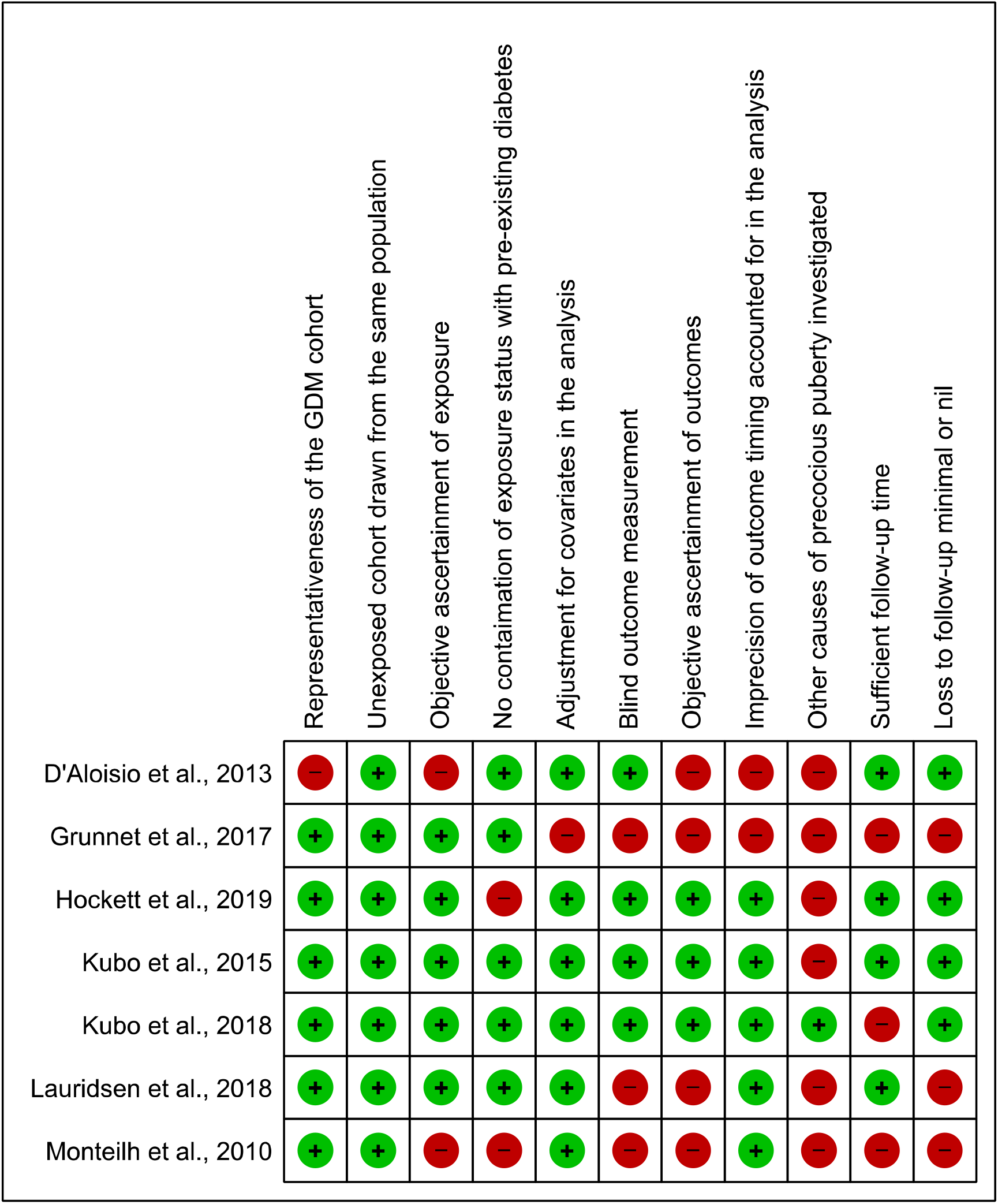
Assessment of risk of bias.

Outcome measurements were performed by research staff in four studies (25-28); three of them specifically reported utilization of recommended methods to measure outcomes, such as orchidometer use for the assessment of testicular size, breast palpation for accurate assessment of the stage of breast development, and computational modelling [Superimposition by Translation And Rotation (SITAR] of longitudinal height measurements for PHV and age at PHV (26-28). Outcomes were recorded only during a series of pre-defined observation times prohibiting the capture of precise pubertal timing, but four studies performed interval censoring to account for this in their analysis (27-30). Notably, two studies recorded Tanner stage in less than 80% of the offspring (29,30), suggesting a possibility of dropout bias.

### Association between maternal gestational diabetes and pubertal timing in girls

Results of the primary studies reviewing the association between maternal GDM and pubertal onset in girls, as indicated by age at menarche, pubarche and thelarche, are given in **Table 2**.

#### Pubic hair development/Pubarche

There was an inconsistent association between maternal GDM and pubarche in girls based on the four primary articles that studied this association. Lauridsen et al. (29) reported an earlier age at attainment of all pubic hair stages in girls of mothers with GDM ranging between 1.6 to 6.0 months after adjustment [adjusted mean monthly difference in PH2: −4.8 (95%CI −7.7, −2.0); PH3: −2.2 (95%CI −4.4, 0); PH4 −1.6 (95%CI −4.8, 1.6); PH5: −6.0 (95%CI −10.8, −1.2)] (**Table 2**).Three studies considered pubertal Tanner stages of ≥PH2 as an outcome.(25,27,28) Grunnet et. Al. (25)**Error! Bookmark not defined**. reported an increase of 51% in age-adjusted odds for reaching ≥PH2 in girls born to mothers with GDM [adjusted OR: 1.51 (95%CI 0.90, 2.55)] (**Table 2**). Kubo et al. conducted two studies in 2016 (27) and 2018 (28)**Error! Bookmark not defined**. using datasets derived from the same database assessing the hazard ratio to reach ≥PH2 for girls of mothers with GDM compared to controls. Both studies did not find a significant association [2015 study adjusted HR: 1.04 (95%CI 0.92, 1.17) and 2018 study adjusted HR: 1.24 (95%CI 0.79, 1.94)] (**Table 2**). When accounting for interaction between maternal pre-gravid BMI and GDM, there was a 3-fold increased hazard of Tanner stage ≥PH2 among girls born to mothers with GDM and a pregravid BMI ≥25 compared to mothers without GDM and a pregravid BMI <25 [adjusted HR: 2.97 (95%CI 1.52, 5.83)].

#### Breast Development /Thelarche

The same four studies that studied the association between pubarche and maternal GDM also studied the association between breast development and GDM (25,27-29). Lauridsen et al. (29)**Error! Bookmark not defined**. did not report significant differences in the mean age at Tanner breast stages 2-5 in girls born to mothers with and without GDM, but the direction of effect size suggest a lower age for all Tanner stages among girls born to mothers with GDM [adjusted mean monthly difference in B2: −4.6 (95%CI −10.1, 1.0); B3: - 1.9 (95%CI −5.0, 1.2); B4: −0.5 (95%CI −3.2, 2.4); B5: −1.8 (95%CI −7.9, 4.3)] (**Table 2**). Grunnet et al. (25) showed twice the age adjusted odds of ≥B2 among girls born to GDM mothers compared to controls [age adjusted OR: 1.99 (95%CI 1.18,3.34)] (**Table 2**); however, once adjusted for offspring BMI, the significance in this association was no longer evident. There was no significant association between maternal GDM and offspring age at thelarche in the 2015 and 2018 studies conducted by Kubo et al [adjusted HR: 0.85 (0.54-1.35) and 1.06 (0.95-1.18) respectively] (27,28) (**Table 2**).

#### Age at peak height velocity

Hockett et al. (26) examined the association between maternal GDM and pubertal timing in the daughters as reflected by growth parameters including peak height velocity (PHV) and age at PHV (APHV). APHV was 10.85 years in girls born to mothers with GDM and 11.12 years in girls born to mothers without GDM, with overlapping confidence intervals (**Table 2**). Using a log-logistic accelerated failure time model, daughters born to mothers with GDM had a 10% higher ethnicity-adjusted height velocity than girls born to mothers without GDM (**Table 2**).

#### Menarche

Maternal GDM seemed to be associated with earlier age at menarche but the evidence is inconsistent. D’Aloisio et al. (24) found that girls born to mothers without pre-gestational or gestational diabetes had no increased risk of earlier (≤10 and 11 years) or later age at menarche (14 and ≥15 years) in comparison to an arbitrary defined reference age of 12-13 years after adjusting for birth decade, ethnicity and family income (**Table 2**). By contrast, girls born to mothers with pregnancy hyperglycemia had a significantly higher risk of earlier menarche (≤10 years) [Adjusted RR: 1.47(95%CI 1.01, 2.16)]. In keeping with these findings, Lauridsen et al. (29) report a significant earlier onset of menarche by 2.5 months in girls born to mothers with GDM compared to mothers without diabetes [adjusted mean monthly difference: - 2.5 (95%CI −4.9, 0)] **(Table 2)**.

### Association between maternal gestational diabetes and pubertal timing in boys

Results of the primary studies reviewing the association between maternal GDM and pubertal onset exclusively among boys indicated by age at spermarche, pubarche and genital development are shown in **Table 3**.

#### Pubic hair development (Pubarche)

Three studies evaluated maternal GDM and its association with pubarche in their sons (**Error! Bookmark not defined**.,**Error! Bookmark not defined**.,**Error! Bookmark not defined**.). Grunnet et al. (25) reported a non-significant 74% increase in the odds of having reached tanner stage ≥PH2 among boys born to mothers with GDM compared to those born to mothers without GDM after adjustment for age [adjusted OR: 1.74 (95%CI 0.92, 3.28)] (**Table 3**). Lauridsen et al. (29) did not find a significant association between maternal GDM and age at pubic hair stages PH2-5, however, they reported trends to earlier ages in GDM boys [Adjusted mean monthly difference for PH2: −1.4 (95%CI −5.3, 2.4); PH3: −1.3 (95%CI −4.6, 1.9); PH4: −0.8 (95%CI −3.4, 1.6); PH5: −1.7 (95%CI −4.7, 1.3)] (**Table 3**). Monteilh et al. (30) performed a step-wise inclusion of covariates based on statistical significance to predict age at transition into stages PH2-4. GDM was not included in the analysis for transition into stages PH2 and PH4 due to statistical insignificance at the predictor selection stage of the analysis^**Error! Bookmark not defined**.^. In the model predicting transition to stage PH3, GDM was included as a predictor along with either offspring BMI or height and weight anthropometrics measures separately recorded at age 8. In the model with BMI, boys born to GDM exposed mothers showed 2-month advancement in the age at transition to PH3 (**Table 3**). Median age of transition to PH3 was 12.6 (95%CI 12.4, 12.7) for boys born to mothers with GDM compared to the entire cohort’s median age of 12.8 (95%CI 12.7, 12.8). In the model with height and weight anthropometrics instead of BMI, median age of transition to PH3 for boys born to mothers with GDM was 12.8 [95%CI 12.6, 13.0] compared to the entire cohort’s median age of transition to PH3 13.0 (95%CI 12.8, 13.1).

#### Genital development and testicular volume

There was no significant association between maternal GDM and the age at onset of male genital development based on the two studies that considered this outcome (25,29). Grunnet et al. (25) reported genital stage ≥G2 in 63 (32.6%) boys of mothers who had GDM and 66 (37.5%) boys of mothers without GDM; after adjusting for their age, they reported an OR of 1.24 (95%CI 0.74, 2.14)**Error! Bookmark not defined**‥ The same study did not report earlier gonadarche (testicular volume ≥ 4mL) in boys born to mothers with GDM [adjusted OR: 0.77 (0.42-1.41)] (**Table 3**). Lauridsen et al. (29) did not find a significant association between maternal GDM and the age at genital stages 2-5 [adjusted mean monthly difference in G2: −0.0 (95%CI −3.8, 3.8); G3: 1.4 (95%CI −1.9, 4.9); G4: 0.5 (85%CI - 2.5, 3.5); G5: 2.6 (95%CI −2.2, 7.4)] (**Table 3**).

#### Age at peak height velocity

Hockett et al. (26) did not find a significant association between the age at PHV among boys and the GDM status of their mothers. However, they reported was a 4% increased PHV among boys born to mothers with GDM compared to boys born to mothers without GDM after adjusting for race/ethnicity (**Table 3**).

#### Spermarche

Lauridsen et al. (29) studied the association between maternal GDM and age at first ejaculation in GDM boys but did not find any significant difference [adjusted mean monthly difference: 0.7 (−2.9, 4.3)] (**Table 3**).

## Discussion

To our knowledge, this is the first systematic review that comprehensively explores the relationship between maternal GDM and pubertal timing; also stratified by offspring gender. Although the current evidence is limited, we noted a subtle trend towards earlier pubertal timing in children exposed to maternal hyperglycemia manifested as GDM *in utero*.

We have included studies that report “maturational events” that are considered to define puberty, i.e. the development of secondary sexual characteristics such as pubic hair, breast (in girls) and penile growth (in boys), growth parameters (such as PHV and age at PHV) and critical events, such as menarche and spermarche.

Although differences in the levels of statistical significance are observed, all of the studies estimate an earlier onset of pubarche in both boys and girls of mothers with GDM. In addition, there was discrepancy in the offspring sex-specific effect of maternal GDM on pubarche. Specifically, Grunnert et al. (25) suggest more pronounced GDM-related odds of pubarche in boys compared to girlswhile Lauridsen et al. (29) report a more pronounced GDM-related precocity of all pubic hair stages in girls compared to boys.

Four studies that examined the onset of breast development (25,27-29) and two studies that examined menarche (24,29) showed variations in the direction, strength and significance of association with maternal GDM. The timing of genital growth and spermarche did not appear to be affected in boys born to mothers with GDM (25,29). One study did collect information on genital development but due to invalidation of longitudinal recording indicated by a significant proportion of boys proposing Tanner stage regression, this outcome was not analysed (30). Growth parameters such as PHV and age at PHV in boys and girls were associated with maternal GDM (26).

Although the present evidence suggests that maternal GDM might be related to early pubertal timing in their offspring, this effect is rather modest or not evident in the full range of pubertal “maturational events”, suggesting a complex interplay between GDM and puberty.

Previous studies have suggested a relationship between maternal GDM and offspring adiposity (31). Adiposity and ‘over-nutrition’ can be considered predictors of pubertal timing and principal determinants for the initiation and maintenance of pubertal maturational events (32), hence, the association between maternal GDM and offspring pubertal timing could be mediated by offspring adiposity and pre-adolescence BMI. This is supported by the analysis by Hockett et al. (26), in which the association between maternal GDM and age at PHV is attenuated by adjustment for offspring BMI z-score.

Several studies have suggested a negative association between pre-pregnancy BMI and timing of puberty (33,34). High pre-pregnancy BMI is an established risk factor of GDM (35); however, considering the available studies it is difficult to dissect the effects of GDM and pre-pregnancy BMI on offspring pubertal timing. Furthermore, an U-shaped association between age at menarche and future risk of GDM has been established (36). Therefore, it is plausible that a synergistic effect exists between the intrauterine effect of hyperglycemia on pubertal timing in the offspring and the genetic influence of earlier maternal age at menarche. In addition to the already explored factors adjusted for in various studies, several other factors such as birthweight (both higher and lower) (37,38), exogenous exposure to endocrine-disrupting chemicals such as phthalates, pesticides and bisphenol A in the mother-offspring home environment (39,40) could have confounded this association. The same applies to leptin, which largely correlates with body fat content. Higher plasma leptin levels have been documented in GDM (41) and may contribute to gestational programming of offspring obesity as leptin is regarded as a permissive signal for puberty initiation (42).

Trends towards earlier pubarche is probably one of the most consistent precocities of all puberty parameters assessed by the studies analysed in this review. It is important to note that the rise of adrenal androgen production in late childhood contributes to the development of pubic (and axillary) hair, an event known as adrenarche (43,44). Adrenarche is a phenomenon currently not well understood, but not related and in fact strictly independent of gonadarche. As adrenarche and gonadarche frequently overlap, it is clinically not possible to distinguish if pubarche is caused by adrenal or testicular androgens in boys, however, it is likely that pubic hair develops as a consequence of adrenal androgen action in girls. Premature adrenarche has been traditionally regarded as benign variant of normal ‘puberty’, however, there is some evidence suggesting that children with premature adrenarche have metabolic dysfunction, in particular abnormal glucose metabolism (45).

To assess the dynamics of pubertal development accurately is difficult, both in the individual clinical setting but even more so based on observational studies. Tanner staging is an unequivocally accepted clinical tool to assess pubertal milestones (46), but prone to inter-observer differences (47,48) and over/under estimation of Tanner staging frequently occurs when being self-reported (49). An LHRH stimulation test as an outcome measure, which would assess the HPG axis most accurately, would aid in objectification as well as differentiation of central and peripheral causes for advancement in pubertal timing (50), albeit difficult to perform in larger study populations due to invasiveness, logistics and cost implications. Rare underlying sinister pathologies, such as sex steroid producing tumours or hypothalamic abnormalities, can affect pubertal timing, however, were only systematically excluded in one of the studies (28).

The findings of the present review should be interpreted in the context of its limitations. One of them was the wide variation in the sample sizes of the included studies. However, it should be noted that no correlation was observed between the sample size and the magnitude or significance of effect estimates. Two pairs of derived their cohorts from the same databases (25,27-29), suggesting a possible overlap of the participants between these pairs of studies.

The summary measures were widely heterogeneous across all of the studies, preventing any meaningful attempt to statistically pool the results. The interval spanned between subsequent observations of Tanner stages or anthropometrics varied across the included longitudinal studies, ranging between 6 months and 1.5 years. Also, there was a high percentage of children who did not agree to report their Tanner stage, which could bias the effect estimates as previous studies report an association between Tanner stage of children and their agreement to have it recorded (51). Therefore, interval and informative censoring embedded in the observational nature of the included studies were potential limitations in accurately discerning the association between maternal GDM and pubertal timing of children.

In order to strengthen the evidence base for the association between maternal GDM and pubertal timing, large-scale prospective cohort studies should be conducted, ideally with objective biochemical tests to measure pubertal timing, standardized approaches in diagnosing GDM and recording of wide range of confounders at baseline. Future research is needed to understand the biological link between the maternal-fetal endocrine system. This can help in the identification of potential interventions to limit the progression of a potential transgenerational continuum of endocrine disturbance and adverse effects on metabolic health.

## Data Availability

Data sharing is not applicable to this article as no datasets were generated or analyzed during the current study.

## Acknowledgements

The authors would like to thank Susan Bayliss, information specialist at the University of Birmingham, for her input into our search strategy.

## References

1. Patton GC, Viner R. Pubertal transitions in health. Lancet. 2007;369(9567):1130–1139.

2. Tanner JM. Trend towards earlier menarche in London, Olso, Copenhagen, the Netherlands and Hungary. Nature. 1973;243(5402):95–96.

3. Sorensen K, Mouritsen A, Aksglaede L, Hagen CP, Mogensen SS, Juul A. Recent secular trends in pubertal timing: implications for evaluation and diagnosis of precocious puberty. Horm Res Paediatr. 2012;77(3):137–145.

4. Marshall WA, Tanner JM. Variations in the pattern of pubertal changes in boys. Arch Dis Child. 1970;45(239):13–23.

5. Marshall WA, Tanner JM. Variations in pattern of pubertal changes in girls. Arch Dis Child. 1969;44(235):291–303.

6. Mendle J, Turkheimer E, Emery RE. Detrimental Psychological Outcomes Associated with Early Pubertal Timing in Adolescent Girls. Dev Rev. 2007;27(2):151–171.

7. Copeland W, Shanahan L, Miller S, Costello EJ, Angold A, Maughan B. Outcomes of early pubertal timing in young women: a prospective population-based study. Am J Psychiatry. 2010;167(10):1218–1225.

8. Day FR, Elks CE, Murray A, Ong KK, Perry JRB. Puberty timing associated with diabetes, cardiovascular disease and also diverse health outcomes in men and women: the UK Biobank study. Sci Rep-Uk. 2015;5.

9. Busch AS, Højgaard B, Hagen CP, Teilmann G. Obesity is associated with earlier pubertal onset in boys. The Journal of Clinical Endocrinology & Metabolism. 2019.

10. Feig DS, Hwee J, Shah BR, Booth GL, Bierman AS, Lipscombe LL. Trends in incidence of diabetes in pregnancy and serious perinatal outcomes: a large, population-based study in Ontario, Canada, 1996-2010. Diabetes Care. 2014;37(6):1590–1596.

11. Joanna briggs institute reviewers’ manual 2014 edition. Institute Joanna Briggs;2014.

12. Stang A. Critical evaluation of the Newcastle-Ottawa scale for the assessment of the quality of nonrandomized studies in meta-analyses. Eur J Epidemiol. 2010;25(9):603–605.

13. Kubo A, Kushi LH, Laurent CA, Greenspan LC, Quesenberry CP, Ferrara A. Maternal pregravid obesity accelerates the timing of pubertal onset in daughters. Diabetes. 2015;64(SUPPL. 1):A427.

14. Hockett CW, Bedrick E, Zeitler P, Crume TL, Daniels SR, Dabelea D. Exposure to Gestational Diabetes Mellitus (GDM) Is Associated with Earlier Pubertal Timing in the Offspring: The EPOCH Study. Diabetes. 2016;65:A42–A42.

15. Hunt S, Hellwig JP. Maternal Obesity, Gestational Diabetes, and Early-Onset Puberty. Nursing for Women’s Health. 2016;20(4):354–354.

16. Brickman W, Catalano P, Clayton PE, et al. Gestational diabetes (GDM) and childhood disorders of glucose metabolism-hyperglycemia and adverse pregnancy outcome follow-up study (HAPO FUS). Diabetes. 2018;67(Supplement 1):A30.

17. Li S, Zhu Y, Yeung E, et al. Offspring risk of obesity in childhood, adolescence and adulthood in relation to gestational diabetes mellitus: A sex-specific association. International Journal of Epidemiology. 2017;46(5):1533–1541.

18. Page KA, Romero A, Buchanan TA, Xiang AH. Gestational diabetes mellitus, maternal obesity, and adiposity in offspring. The Journal of pediatrics. 2014;164(4):807–810.

19. Persson I, Ahlsson F, Ewald U, et al. Influence of perinatal factors on the onset of puberty in boys and girls: implications for interpretation of link with risk of long term diseases. American journal of epidemiology. 1999;150(7):747–755.

20. Ibanez L, Castell C, Tresserras R, Potau N. Increased prevalence of type 2 diabetes mellitus and impaired glucose tolerance in first-degree relatives of girls with a history of precocious pubarche. Clinical endocrinology. 1999;51(4):395–401.

21. Tertti K, Toppari J, Virtanen HE, Sadov S, Ronnemaa T. Metformin treatment does not affect testicular size in offspring born to mothers with gestational diabetes. Review of diabetic studies. 2016;13(1):59–65.

22. Hockett CW, Harrall KK, Moore BF, et al. Persistent effects of in utero overnutrition on offspring adiposity: the Exploring Perinatal Outcomes among Children (EPOCH) study. Diabetologia. 2019.

23. Davis JN, Gunderson EP, Gyllenhammer LE, Goran MI. Impact of gestational diabetes mellitus on pubertal changes in adiposity and metabolic profiles in Latino offspring. The Journal of pediatrics. 2013;162(4):741–745.

24. D’Aloisio AA, DeRoo LA, Baird DD, Weinberg CR, Sandler DP. Prenatal and infant exposures and age at menarche. Epidemiology (Cambridge, Mass). 2013;24(2):277–284.

25. Grunnet LG, Hansen S, Hjort L, et al. Adiposity, Dysmetabolic Traits, and Earlier Onset of Female Puberty in Adolescent Offspring of Women With Gestational Diabetes Mellitus: A Clinical Study Within the Danish National Birth Cohort. Diabetes care. 2017;40(12):1746–1755.

26. Hockett CW, Bedrick EJ, Zeitler P, Crume TL, Daniels S, Dabelea D. Exposure to Diabetes in Utero Is Associated with Earlier Pubertal Timing and Faster Pubertal Growth in the Offspring: The EPOCH Study. Journal of Pediatrics. 2019;206:105–112.

27. Kubo A, Ferrara A, Laurent CA, et al. Associations Between Maternal Pregravid Obesity and Gestational Diabetes and the Timing of Pubarche in Daughters. American journal of epidemiology. 2016;184(1):7–14.

28. Kubo A, Deardorff J, Laurent CA, et al. Associations Between Maternal Obesity and Pregnancy Hyperglycemia and Timing of Puberty Onset in Adolescent Girls: A Population-Based Study. American journal of epidemiology. 2018;187(7):1362–1369.

29. Lauridsen LLB, Arendt LH, Ernst A, et al. Maternal diabetes mellitus and timing of pubertal development in daughters and sons: a nationwide cohort study. Fertility and sterility. 2018;110(1):35–44.

30. Monteilh C, Kieszak S, Flanders WD, et al. Timing of maturation and predictors of Tanner stage transitions in boys enrolled in a contemporary British cohort. Paediatric and perinatal epidemiology. 2011;25(1):75–87.

31. Logan KM, Gale C, Hyde MJ, Santhakumaran S, Modi N. Diabetes in pregnancy and infant adiposity: systematic review and meta-analysis. Arch Dis Child Fetal Neonatal Ed. 2017;102(1):F65–F72.

32. Li WY, Liu Q, Deng X, Chen YW, Liu SD, Story M. Association between Obesity and Puberty Timing: A Systematic Review and Meta-Analysis. Int J Env Res Pub He. 2017;14(10).

33. Brix N, Ernst A, Lauridsen LLB, et al. Maternal pre-pregnancy body mass index, smoking in pregnancy, and alcohol intake in pregnancy in relation to pubertal timing in the children. BMC Pediatr. 2019;19(1):338.

34. Brix N, Ernst A, Lauridsen LLB, et al. Maternal pre-pregnancy obesity and timing of puberty in sons and daughters: a population-based cohort study. Int J Epidemiol. 2019;48(5):1684–1694.

35. Torloni MR, Betrán AP, Horta BL, et al. Prepregnancy BMI and the risk of gestational diabetes: a systematic review of the literature with meta-analysis. Obesity Reviews. 2009;10(2):194–203.

36. Petry CJ, Ong KK, Hughes IA, Acerini CL, Dunger DB. The association between age at menarche and later risk of gestational diabetes is mediated by insulin resistance. Acta Diabetol. 2018;55(8):853–859.

37. Deng X, Li WY, Luo Y, Liu SD, Wen Y, Liu Q. Association between Small Fetuses and Puberty Timing: A Systematic Review and Meta-Analysis. Int J Env Res Pub He. 2017;14(11).

38. Wang Y, Dinse GE, Rogan WJ. Birth weight, early weight gain and pubertal maturation: a longitudinal study. Pediatr Obes. 2012;7(2):101–109.

39. Ehrlich S, Lambers D, Baccarelli A, Khoury J, Macaluso M, Ho SM. Endocrine Disruptors: A Potential Risk Factor for Gestational Diabetes Mellitus. Am J Perinatol. 2016;33(13):1313–1318.

40. Fisher MM, Eugster EA. What is in our environment that effects puberty? Reprod Toxicol. 2014;44:7–14.

41. Kautzky-Willer A, Pacini G, Tura A, et al. Increased plasma leptin in gestational diabetes. Diabetologia. 2001;44(2):164–172.

42. Elias CF. Leptin action in pubertal development: recent advances and unanswered questions. Trends Endocrinol Metab. 2012;23(1):9–15.

43. Idkowiak J, Lavery GG, Dhir V, et al. Premature adrenarche: novel lessons from early onset androgen excess. Eur J Endocrinol. 2011;165(2):189–207.

44. Auchus RJ, Rainey WE. Adrenarche - physiology, biochemistry and human disease. Clin Endocrinol (Oxf). 2004;60(3):288–296.

45. Liimatta J, Utriainen P, Laitinen T, Voutilainen R, Jaaskelainen J. Cardiometabolic Risk Profile Among Young Adult Females With a History of Premature Adrenarche. J Endocr Soc. 2019;3(10):1771–1783.

46. Mendle J, Beltz AM, Carter R, Dorn LD. Understanding Puberty and Its Measurement: Ideas for Research in a New Generation. J Res Adolesc. 2019;29(1):82–95.

47. Slora EJ, Bocian AB, Herman-Giddens ME, et al. Assessing Inter-Rater Reliability (IRR) of Tanner Staging and Orchidometer Use with Boys: A Study from PROS. Journal of Pediatric Endocrinology & Metabolism. 2009;22(4):291–299.

48. Carlsen E, Andersen AG, Buchreitz L, et al. Inter-observer variation in the results of the clinical andrological examination including estimation of testicular size. Int J Androl. 2000;23(4):248–253.

49. Chavarro JE, Watkins DJ, Afeiche MC, et al. Validity of Self-Assessed Sexual Maturation Against Physician Assessments and Hormone Levels. J Pediatr. 2017;186:172-178.e173.

50. Tirumuru SS, Arya P, Latthe P, Kirk J. Understanding precocious puberty in girls. The Obstetrician & Gynaecologist. 2012;14(2):121–129.

51. Baird J, Walker I, Smith C, Inskip H. Review of methods for determining pubertal status and age of onset of puberty in cohort and longitudinal studies.: MRC Lifecourse Epidemiology Unit, University of Southampton;2017.

